# Personalized Insights Derived from Wearable Device Data and Large Language Models to Improve Well-Being

**DOI:** 10.64898/2026.03.03.26347299

**Authors:** Kai He, Yu Fang, Elena Frank, Chunyu Li, Amy Bohnert, Srijan Sen, Meng Wang

## Abstract

Health behaviors such as physical activity and sleep affect mental health, but the effect of each health behavior varies substantially across individuals, limiting the usefulness of generic behavioral recommendations. We collected one year of continuous wearable and ecological momentary assessment data from 3,139 participants in the Intern Health Study (2018–2023), and examined individual-level associations between wearable-derived features and mood across the internship year. The behaviors associated with mood were highly heterogeneous between individuals: the two most prevalent drivers of mood were wake-up time (the strongest driver for 34.0% of subjects) and step count (10.6% of subjects). The correlation directionality remained largely stable despite fluctuations in strength. Interestingly, 20.3% of subjects showed no significant correlations. These findings highlight the limitations of population-level recommendations and the critical need for personalized, data-driven approaches to mental health assessment and intervention. To translate these personalized insights into actionable support, we developed MoodDriver, a large language models (LLM)-powered system that generates tailored feedback emails based on each participant’s behavioral and physiological patterns. This work demonstrates the feasibility of combining digital phenotyping with large language models to advance precision digital mental health for high-risk populations.

## Introduction

With growing evidence that health behaviors, such as sleep and physical activity, affect depression, anxiety, and emotional stability^1–5^, there has been growing interest in lifestyle interventions targeting health behaviors. Recent studies have identified substantial interindividual variability in behavioral responses to lifestyle interventions^6–8^, indicating that individuals do not benefit equally from changes in activity or sleep patterns. However, there remains limited evidence specifying which behaviors are most critical for optimizing mental health in which individuals and contexts. Consequently, most lifestyle intervention and health behavior recommendations remain broad and generic rather than personalized or adaptive, limiting their effectiveness.

Digital health technologies have rapidly transformed mental health research and care by enabling scalable and continuous data collection through smartphones and wearable devices^8–13^. These technologies can continuously and passively monitor physiological and behavioral factors such as heart rate, step count, and sleep duration and timing, providing rich, longitudinal data under naturalistic environments. Medical internship, the first-year of professional physician training, involves long work hours, frequent night shifts and challenging clinical situations. With the onset of internship, health behaviors such as step count, sleep duration and consistency and circadian alignment worsen and, correspondingly, depression rates increase 4-5 fold^14^. We analyzed data from 3,139 medical interns enrolled in the longitudinal Intern Health Study (IHS)^15^ between 2018 and 2023 to examine individual-level associations between wearable features and mood dynamics. We further developed an LLM-powered system, MoodDriver, to generate tailored feedback emails that inform participants how their behaviors relate to mood.

## Results

### Heterogeneous patterns of mood-feature correlations

We utilized Spearman correlation to assess the associations between wearable-derived features and daily mood scores for each participant across the full internship year. To characterize the correlation patterns, we grouped each intern–feature pair into three categories based on correlation significance and direction: non-significant correlation (Non-Sig Cor), significant positive correlation (Sig-Pos Cor), and significant negative correlation (Sig-Neg Cor).

The correlation heatmap (Fig. 1a) reveals highly heterogeneous patterns of mood–feature relationships across both individuals and feature types. Each feature evaluated was both Sig-Pos Cor and Sig-Neg Cor associations with mood for some individuals, suggesting substantial interindividual variability. For instance, higher step count was significantly positively associated with mood in 26.9% of subjects and significantly negatively associated with mood in 5.8% of subjects. Across the cohort, we observed marked heterogeneity in the number and direction of significant mood–feature associations (Fig. 1b); 59.4% of interns had more than two significant mood correlates, while 20.3% exhibited no significant mood–feature associations. Among sleep-related features, wake-up time showed the highest proportion of Sig-Pos Cor (49.0%), followed by bedtime (31.8%) and sleep duration (31.5%). The proportions of interns in each correlation category across all features are shown in Fig. 1c. We further examined the distribution of the strongest drivers among these subjects. For positive associations, wake-up time was the most prominent driver (34.0%), followed by step count (10.6%), active minutes (10.0%), bedtime (8.8%), sleep duration (6.7%), and resting heart rate (3.4%). In contrast, for negative associations, resting heart rate was the most influential driver (15.6%), followed by active minutes (5.1%), step count (4.1%), wake-up time (0.9%), sleep duration (0.9%), and bedtime (0.7%). Together, these findings demonstrate pronounced heterogeneity in mood–feature correlation patterns across individuals.

**Fig 1.**
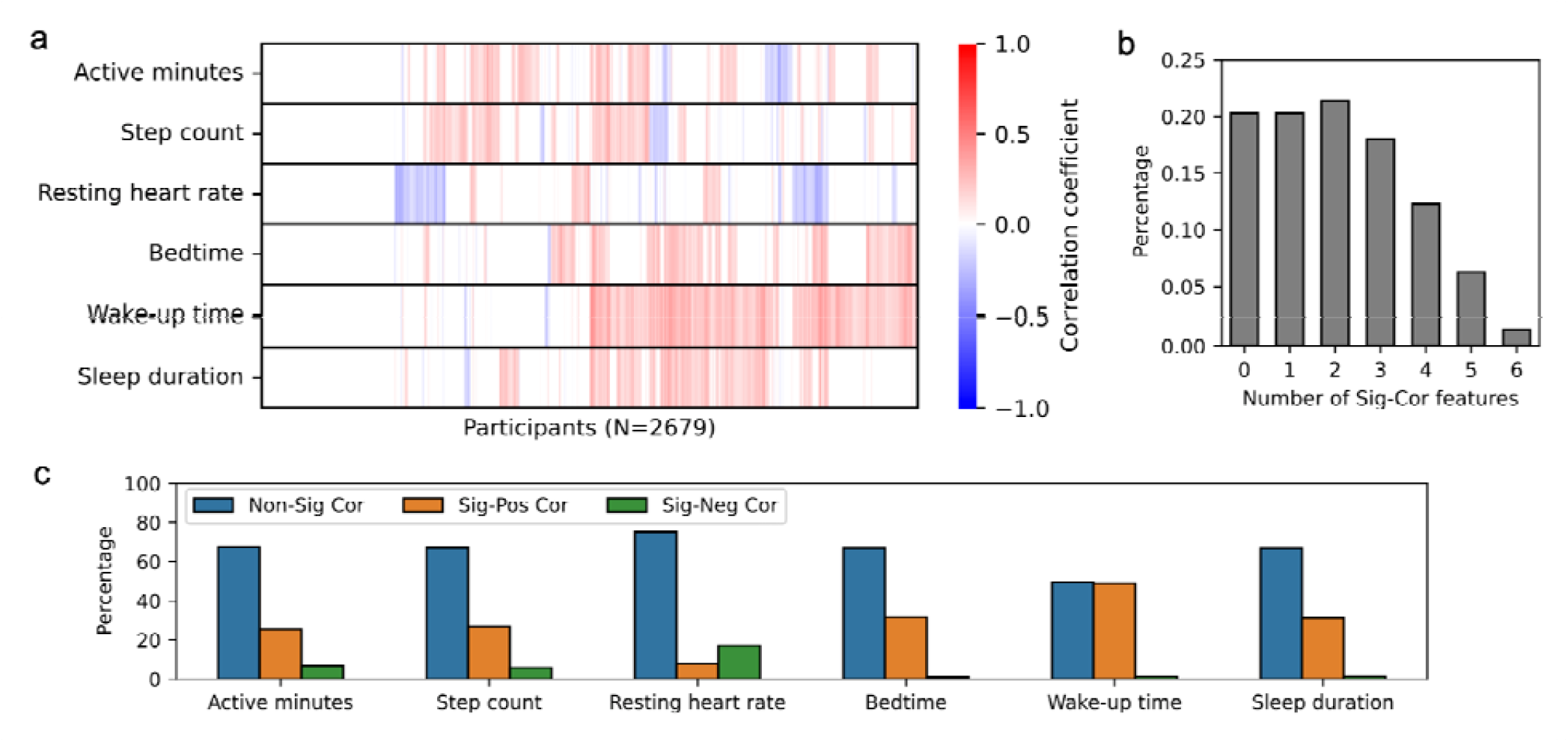
Heterogeneity of mood-feature correlations across participants and wearable-derived features. (a) Heatmap of Spearman correlations between mood scores and wearable-derived features across interns during the internship year. Only participants meeting the non-missing-value criteria were included (see Methods). Correlation coefficients () with were set to zero for visualization. Based on correlation direction and significance, each association was categorized as significant positive (Sig-Pos Cor;), significant negative (Sig-Neg Cor;), or non-significant (Non-Sig Cor;). (b) Distribution of interns by the number of significant mood–feature associations. (c) Distribution of interns across correlation categories (Sig-Pos Cor, Sig-Neg Cor, Non-Sig Cor) for each wearable-derived feature.

### Temporal dynamics of mood-feature correlations

To examine how mood–feature associations evolved over time, we investigated their temporal dynamics across the four quarters of the internship year. For each quarter, we performed the same Spearman correlation analysis and categorized each intern–feature pair into one of three groups: Sig-Pos Cor, Sig-Neg Cor, or Non-Sig Cor. The transitions among these categories across consecutive quarters for all features are shown in Fig. 2a–f.

**Fig 2.**
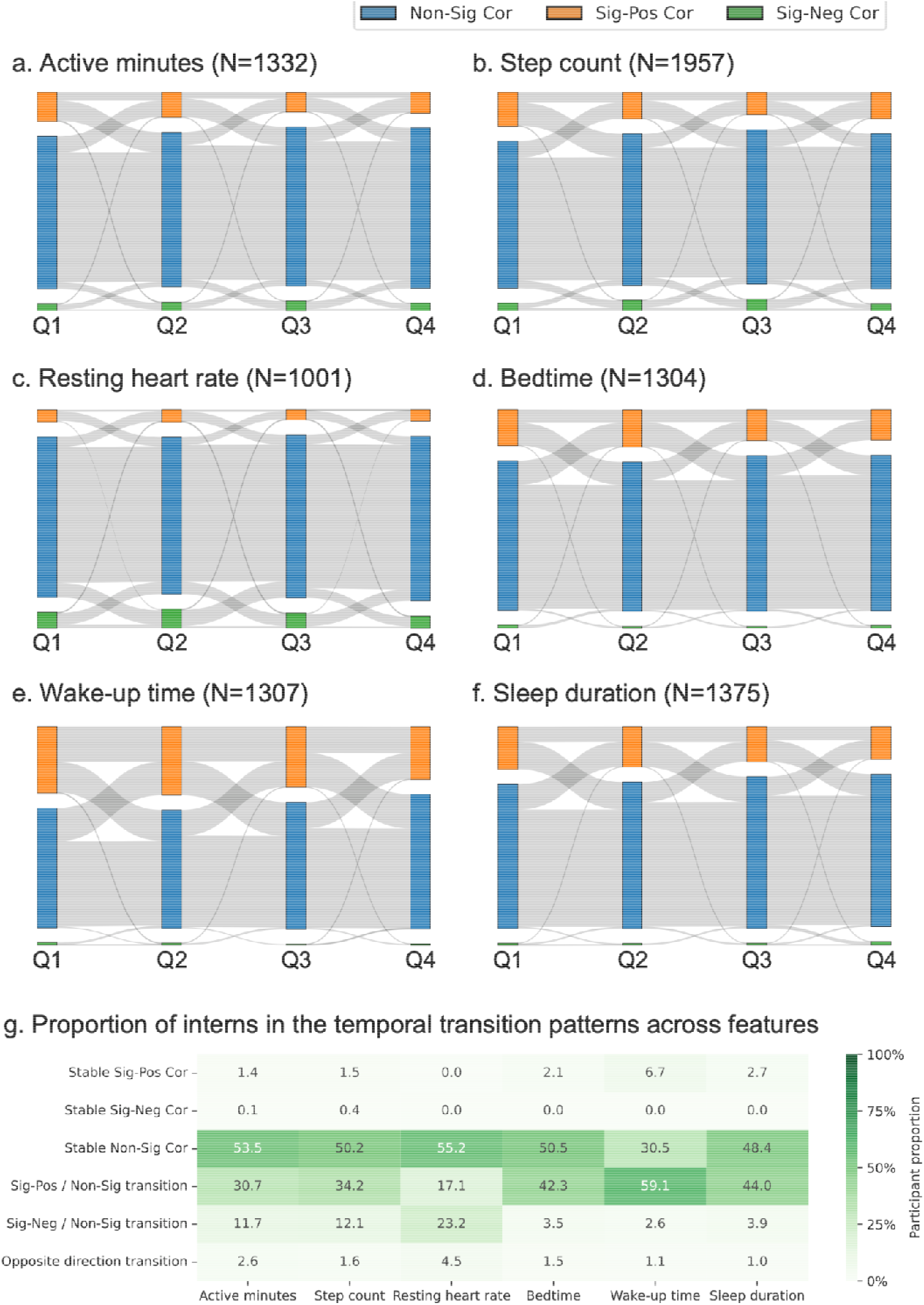
Temporal dynamics of mood–feature correlations across four quarters of the internship year. (a–f) Sankey plots showing the number of participants transitioning among the three correlation categories for each wearable-derived feature across quarters (Q1–Q4). Spearman correlations between mood and each feature were computed within each quarter, and correlation categories were assigned according to the same criteria. Only participants meeting the non-missing-value criteria were included (see Methods). (g) Heatmap showing the proportion of interns in the selected temporal transition patterns across features. The transition patterns include: (i) Stable Sig-Pos Cor—significant positive correlations in all four quarters; (ii) Stable Sig-Neg Cor— significant negative correlations in all four quarters; (iii) Stable Non-Sig Cor—non-significant correlations in all four quarters; (iv) transitions between Sig-Neg Cor and Non-Sig Cor; (v) transitions between Sig-Pos Cor and Non-Sig Cor; and (vi) opposite correlation direction transition.

To further characterize individual-level temporal patterns, we grouped transition trajectories in six representative patterns for each feature: (i) Stable Sig-Pos Cor – significant positive correlations in all four quarters; (ii) Stable Sig-Neg Cor – significant negative correlations in all four quarters; (iii) Stable Non-Sig Cor – non-significant correlations in all four quarters; (iv) transitions between Sig-Pos Cor and Non-Sig Cor; (v) transitions between Sig-Neg Cor and Non-Sig Cor; and (vi) transitions between Sig-Pos Cor and Sig-Neg Cor. Across all features, only a small minority of participants fell into the Stable Sig-Pos Cor and Stable Sig-Neg Cor trajectories (0 – 6.7%), while a substantial portion of the sample (30.5 – 55.2%) fell into the Stable Non-Sig Cor trajectory. The remaining 44.8 – 62.8% of participants demonstrated temporal variation in their correlations, indicating that stable, consistently significant correlations were uncommon. Among those with changing patterns, transitions between Sig-Neg Cor ↔ Non-Sig Cor and Sig-Pos Cor ↔ Non-Sig Cor were the most common (40.3-61.7 %), indicating fluctuations in the strength but not the direction of associations. These findings reveal that, while the directionality of associations tended to remain consistent, the strength of these relationships varied over time.

### Variations in effect sizes among significant mood-feature relationships

To quantify effect sizes, we applied simple linear regression for each feature (Sig-Pos Cor or Sig-Neg Cor), regressing mood scores on the corresponding feature values (Fig. 3). The slope from the linear regression quantifies the extent to which changes in a feature are associated with corresponding changes in mood scores. Active minutes had slopes of 0.019 (±0.051) for Sig-Pos Cor and −0.028 (±0.057) for Sig-Neg Cor. Similar bidirectional patterns were observed for step count (0.114 ± 1.035; −0.186 ± 1.936) and resting heart rate (0.156 ± 0.280; −0.156 ± 0.258). Sleep-related metrics showed larger variability: wake-up time (0.581 ± 6.351; −1.002 ± 8.532), bedtime (0.413 ± 1.837; −0.442 ± 2.310), and sleep duration (0.365 ± 2.420; −0.452 ± 5.066). Collectively, these results indicate substantial heterogeneity in personalized slopes, with considerable variability even within the same association category. We found that, averaging across subjects, 1 point mood improvement was associated with a 38-minute increase in active minutes, 6.6k additional steps, going to bed 2 hours earlier, waking up 4 hours later, and extending total sleep by 4 hours.

**Fig 3.**
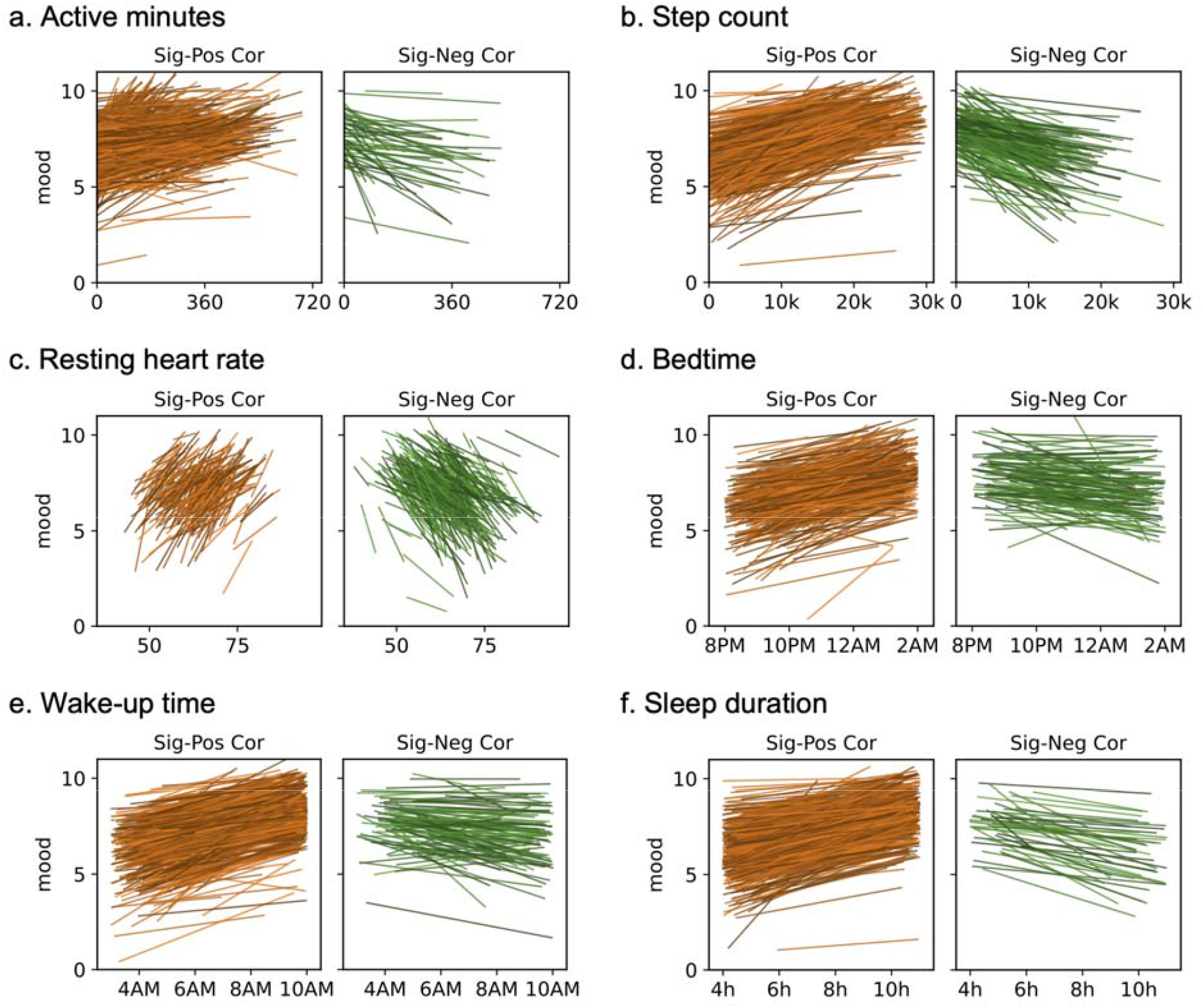
Quantification of individual-level mood–feature relationships. (a–f) Simple linear regression models estimating the relationship between mood scores and each wearable-derived feature among participants in the Sig-Pos Cor and Sig-Neg Cor groups. Results are shown for the first quarter of the internship year. The fitted slopes quantify how changes in each feature are associated with corresponding changes in mood at the individual level. For visualization, line color brightness was randomly assigned to distinguish individual regression lines.

### LLM-derived mood feedback system — MoodDriver

Building on our identified associations, we utilized LLMs to develop a feedback system, MoodDriver, to deliver personalized analyses and feedback to individuals, explaining the unique relationships between their behavior and mood. The overall framework of MoodDriver is illustrated in Fig. 4.

**Fig 4.**
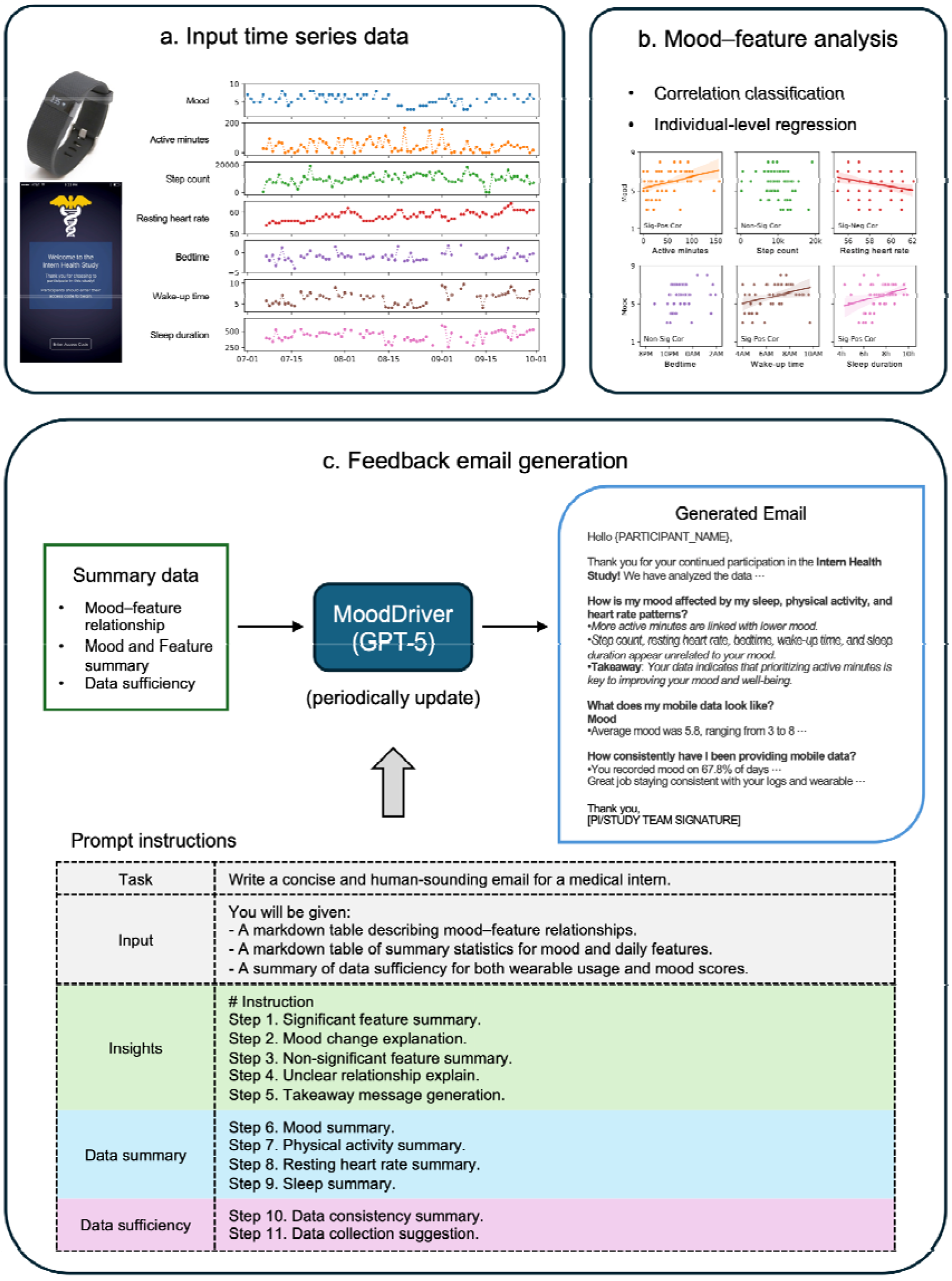
Overview of the LLM-powered mood feedback system (MoodDriver). (a) Example input time-series data of mood scores and wearable-derived features from one intern during the first quarter of internship. (b) Mood–feature analysis for the example intern, including correlation classification and individual-level regression for significant features. (c) MoodDriver framework for generating personalized mood feedback emails, integrating summary statistics and analysis outputs using GPT-5 with task-specific prompt instructions according to the email template.

Utilizing the time series of mood scores and the collected wearable-derived features (active minutes, step count, resting heart rate, bedtime, wake-up time, and sleep duration) as input (Fig. 4a), MoodDriver computes Spearman correlations between mood and each feature, classifying the relationships into Non-Sig Cor, Sig-Pos Cor, or Sig-Neg Cor categories (Fig. 4b). For features with significant correlations, MoodDriver conducts individual-level univariate linear regressions, modeling mood as a function of each feature to estimate the strength of association (Fig. 4b). MoodDriver summarizes three key components: (i) mood–feature relationship, including correlation categories and regression slopes; (ii) the input mood and feature data; and (iii) data sufficiency, measured as the proportion of non-missing values (see Methods). Recognizing the potential for hallucination—a known limitation of generative models^16^, we developed structured prompt instructions and constrained input templates to ensure factual consistency and alignment with the underlying statistical analyses. Leveraging GPT-5 and customized prompt engineering, MoodDriver integrates these analytic outputs into personalized, conversational feedback messages, delivered as emails written in natural language (Fig. 4c, **Supplemental Materials**). Representative messages for eight participants are shown in Fig. 5. These examples include individuals with no significant mood correlates (No drivers), multiple significant correlates (Multiple drivers), and those whose moods were negatively or positively associated with either activity features (active minutes, steps), sleep features (bedtime, wake-up time, sleep duration), or resting heart rate (RHR). Each feedback email begins with a brief acknowledgment and specifies the data range used in the analysis. MoodDriver then delivers personalized insights by addressing three guiding questions. First, in response to “How is my mood affected by my sleep, physical activity, and heart rate patterns?”, MoodDriver summarizes all identified mood–feature relationships as bullet points, such as “More active minutes, later wake-up times, and longer sleep are linked with better mood”, “For each additional hour of active time, your mood score decreases by 0.6 points”, or “For each additional hour of sleep, your mood score decreases by 0.68 points”. These statements are followed by a concise takeaway message that highlights the significant mood drivers for improving emotional well-being. For individuals without significant mood correlates (e.g., Intern 1), MoodDriver explicitly reports the absence of detected associations and suggests that other factors, such as social connection, daylight exposure, or nutrition, may play a more prominent role. Second, in response to “What does my mobile data look like?”, MoodDriver reports summary statistics for mood and wearable-derived features, including mean values and ranges, presented as bullet points. When data are unavailable for a given feature, this is explicitly noted (e.g., “No data available”). Third, in response to “How consistently have I been providing mobile data?”, MoodDriver summarizes data frequency and completeness for mood and sensor-derived features, again using bullet points, regardless of data availability. At the end of the email, MoodDriver encourages users to maintain consistent data collection and highlights areas for improvement, for example: “Great job wearing your device consistently during the day and night; try logging your mood more regularly so we can provide clearer and more reliable insights.”

**Fig 5.**
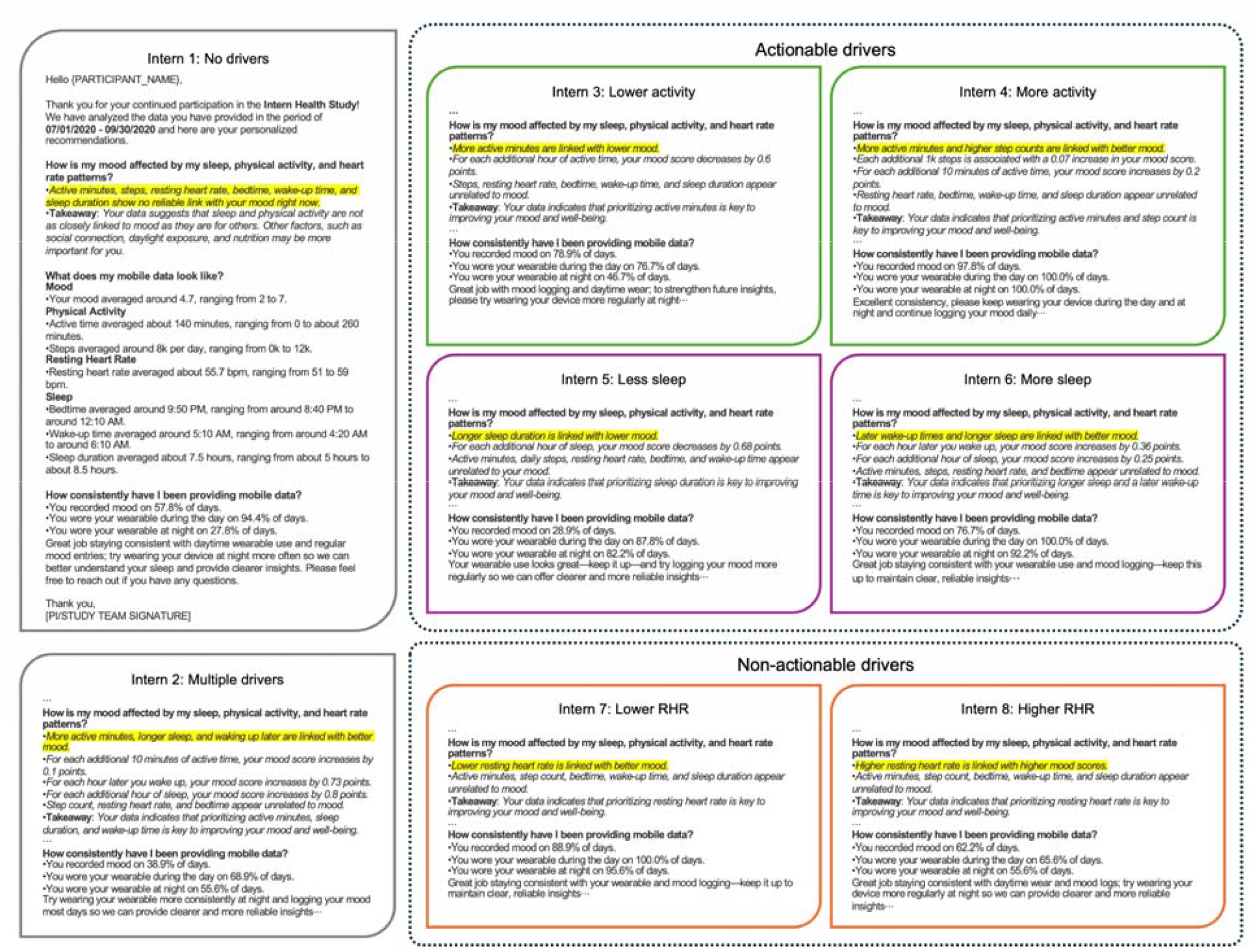
Personalized mood feedback emails generated by MoodDriver for eight interns. Example personalized feedback emails automatically generated by MoodDriver. The eight interns were randomly selected to represent distinct types of mood drivers (significant correlates), including: no drivers, multiple drivers, actionable drivers (less activity, more activity, less sleep, more sleep), and non-actionable drivers (lower resting heart rate [RHR], higher RHR). Each email highlights the individualized interpretation of relationships between mood and wearable-derived features identified for that intern.

## Discussion

In this study, we assessed individual-level associations between smartwatch-derived behavioral and physiological features (activity, sleep, and resting heart rate) and mood dynamics across 3,139 training physicians through their intern year. We found substantial interindividual heterogeneity, with distinct subsets of subjects showing significant positive correlations, significant negative correlations and no significant correlations for all features. While the direction of association often varied between individuals, the direction of associations remained largely stable within individuals, despite fluctuations in strength and significance. Notably, 20.3% of interns exhibited no significant mood correlates, suggesting that a subset of the population is not sensitive to the set of health behaviors assessed in this study.

Previous digital health interventions have typically relied on generic recommendations such as broadly encouraging increased physical activity as universally beneficial strategies^17–19^. However, our findings reveal that the population-level associations mask individual-level heterogeneity. Physical activity, for instance, was associated with improved mood overall, but only 25.5% of participants showed the expected significant positive association, and higher activity correlated with lower mood levels for 6.9% interns. These results highlight the individual variability in behavioral and physiological drivers of emotional well-being and how aggregated population-level recommendations may have limited relevance for a meaningful subset of the population.

Building on these findings, we developed an LLM-powered feedback system, MoodDriver, which translates associations between mobile technology features and mood into tailored, specific and interpretable messages linking behaviors and physiological factors with mood. MoodDriver offers a scalable, data-driven framework for promoting adaptive behavior change through enhancing individuals’ awareness of their behavioral and physiological drivers of mood.

The MoodDriver system advances beyond traditional, population-level messaging by providing tailored, personalized mood feedback grounded in each individual’s data. LLM-based AI agents have recently been applied in digital mental health care, supporting tasks such as mood monitoring, reflective dialogue, cognitive-behavioral guidance, and personalized behavioral coaching^20–24^. Studies have demonstrated the potential of LLM-based AI agents to enhance engagement and deliver supportive interactions, though challenges remain around safety, transparency, and clinical validation. By integrating an LLM directly into a personalized digital phenotyping pipeline, MoodDriver extends this work by linking individual behavioral, physiological data and mood, real-time statistical modeling, and natural-language feedback generation. This approach represents a step toward intelligent, adaptive, and evidence-grounded digital mental health support systems that can scale to broad, real-world populations. In future work, we will assess whether receipt of MoodDriver output improves individual mood and the specific health behaviors that are related to one’s mood.

There are several limitations to our study. First, missing data may have affected statistical power, particularly for participants with irregular wearable use. Second, our analysis was limited to a predefined set of wearable-derived features (activity, sleep, and resting heart rate). Additional features could be added as data availability permits. Incorporating additional contextual data, such as environmental factors, social interactions, or work schedules, could further improve model precision and interpretability. Third, the study primarily utilized data from Fitbit devices; however, the analytical and feedback framework is device-agnostic and can be readily extended to other platforms such as Apple Watch or Garmin. Fourth, this study focused on medical interns, a high-risk population for mood disturbances. While this group provides a valuable model for studying stress-related dynamics, the MoodDriver should be assessed in other populations.

In conclusion, this study demonstrates that mood–behavior relationships are highly individualized and dynamic. By combining continuous wearable data with large language models, our MoodDriver framework translates personalized statistical analyses into interpretable, data-driven feedback that supports self-awareness and mood regulation. This approach offers a scalable path toward precision digital mental health, enabling adaptive and personalized interventions that evolve with each individual’s behavioral and physiological patterns.

## Methods

### Participants

The Intern Health Study (IHS)^15,25,26^ is a US national annual cohort study that follows physicians through their first year of internship. Training physicians starting internship between 2018-2023 were invited to take part in the study. Prior to the start of internship, participants provided consent and completed the initial survey via the MyDataHelps study mobile application and were provided a wrist-worn Fitbit device. Participants received a daily push notification prompting them to complete a single-item mood assessment with the question: ‘On a scale of 1 (lowest) to 10 (highest), how was your mood today?’

For each participant, six daily features were extracted from the Fitbit devices: active minutes, step count, resting heart rate, bedtime, wake time, and sleep duration. A total of 3,139 interns were included in this study (55.9% female; mean age 27.8 ± 2.7 years; see Extended Data Fig. 1 for details of participant inclusion). On average, 262.3 (±111.2) days of wearable data and 103.3 (±100.9) days of mood scores were collected for each participant. The study was approved by the institutional review board at the University of Michigan. All subjects provided informed consent after receiving a complete description of the study. All procedures were conducted in accordance with the Declaration of Helsinki.

### Data Processing

Due to changes in the Fitbit API, the available physical activity (PA) features differed across enrollment cohorts. For participants enrolled between 2018 and 2020, total active minutes were assessed. For participants enrolled between 2021 and 2023, three levels of active minutes were available: lightly active, fairly active, and very active minutes. To harmonize activity measures across cohorts^27^, active minutes for participants enrolled after 2021 were calculated as the sum of fairly active and very active minutes, consistent with the definition of active minutes as activities exceeding 3 metabolic equivalents (METs). We derived bedtime and wake-up time from the main sleep episode recorded by the Fitbit device each day. Total sleep duration was calculated by summing all sleep episodes within the same day.

Outliers were filtered using fixed thresholds (based on the distributions of wearable data in the sample, see Extended Data Fig 2) to mitigate their influence on the linear model fitting on a daily level: active minutes greater than 720, steps greater than 30k, and resting heart rate above 100 were excluded from the analysis. Regular (non-shift) sleep was defined as sleep episodes with bedtimes between 8:00 PM and 2:00 AM, wake-up times between 3:00 AM and 10:00 AM, and sleep durations between 4 and 11 hours. To ensure consistency, only regular sleep-related features were included in subsequent analyses.

### Mood–feature analysis

To quantify the relationships between wearable-derived features and mood, we computed Spearman’s rank correlations (SciPy, Python)^28^ between each feature and mood scores for every participant. Mood–feature associations were calculated both across the full year and quarterly. For the whole-year analysis, subjects were included if they had at least 30 valid mood–feature pairs for each of the six features. In total, 2,679 subjects met the criteria for inclusion. For the quarterly analysis, each wearable-derived feature was evaluated separately to maximize participant inclusion. Participants were included if they had at least 10 valid mood–feature pairs within a given quarter for all quarters. The numbers of participants meeting this criterion were 1,332 for active minutes, 1,957 for step count, 1,001 for resting heart rate, 1,304 for bedtime, 1,307 for wake time, and 1,375 for sleep duration, resulting in a total of 2,191 subjects included in the quarterly analysis. Overall, 3,139 subjects were included in at least one section of the analyses. Based on the correlation results, each feature for each participant was assigned to one of four categories: (i) significant positive correlation (Sig-Pos Cor), (ii) significant negative correlation (Sig-Neg Cor), (iii) non-significant correlation (Non-Sig Cor), or (iv) uncertain (the Spearman correlation could not be computed). For results with available p-values, a p-value threshold of 0.05 was used for classification: correlations with □ < *0.05* were classified as Sig-Pos Cor or Sig-Neg Cor depending on the sign of the correlation coefficient, while those with □ > *0.05* were labeled as Non-Sig Cor. For significant correlations, simple linear regression was performed at the individual level, regressing mood scores on the corresponding feature values. The slope coefficient from each model was extracted to quantify the extent to which changes in the feature were associated with changes in mood. To visualize temporal dynamics in mood– feature associations across quarters, Sankey plots were generated for each feature individually using the Plotly package^29^ in Python.

### Implementation of MoodDriver

For each participant and each quarter, we organized input data for MoodDriver into three components: (i) a mood–feature relationship summary, (ii) a mood and feature data summary, and (iii) data sufficiency indicators. The mood-feature summary reported the assigned association category (Sig-Pos Cor, Sig-Neg Cor, Non-Sig Cor, or uncertain), and the individual-specific slope for features exhibiting significant correlations. For bedtime and wake-up time, summary statistics were calculated only from regular sleep episodes (i.e., non–night-shift sleep periods). When a participant’s sleep behavior indicated a night-shift schedule, bedtime and wake-time minimum and maximum values were set to missing. To characterize data sufficiency, we quantified completeness across three dimensions: daytime data sufficiency, nighttime data sufficiency, and mood score sufficiency. For each category, data sufficiency was classified as high when data were available for more than 45 out of 90 days within a quarter, and as low when data were available for 45 or fewer days. Daytime sufficiency was defined by the minimum availability across active minutes and step count. Nighttime sufficiency was defined by the least availability among resting heart rate, sleep duration, bedtime, and wake time. Mood sufficiency was based on the availability of mood scores. A comprehensive prompt with structured instructions (Fig. 4c) was used to guide GPT-5 in generating personalized mood reports. The full prompt and email template are provided in the Extended Data. All GPT-5 hyperparameters were set to default values.

The MoodDriver email report sent to participants is composed of three main sections: 1) interpretation of mood–feature relationships; 2) summaries of behavioral features and mood states; and 3) data collection suggestions. Feedback emails were generated using first-quarter data from participants. The email reports were produced using the UM GPT API^30^ and Pandas^31^ in Python. To illustrate the range of report types, we selected representative examples reflecting distinct correlation patterns with mood, including: no drivers, multiple drivers, lower activity, more activity, less sleep, more sleep, lower RHR, and higher RHR. For each category, three examples were randomly selected. Full input and output data for all selected cases are provided in the Supplemental Table.

## Data availability

The de-identified data from Intern Health Study that support the findings described here are available from the corresponding author upon reasonable request.

## Code availability

The code for data processing and analysis is available upon reasonable request.

## Acknowledgements

We thank the medical interns who participated in Intern Health Study. K. H., C.L., and M.W. were supported by M.W.’s startup fund from the Institute for Heart and Brain Health. We were all supported by NIH (R01MH101459, U01MH136025). This research was supported in part through computational resources and services provided by Advanced Research Computing at the University of Michigan, Ann Arbor.

## Author contributions

M.W. and S.S. conceived the study and supervised the research. K. H. performed the analysis and implemented the LLMs. K. H., Y.F., E.F., C.L., A.B., S.S., and M.W. discussed the results, wrote the manuscript, and revised it.

