## Supplementary Files for "Personalized Insights Derived from Wearable Device Data and Large Language Models to Improve Well-Being"

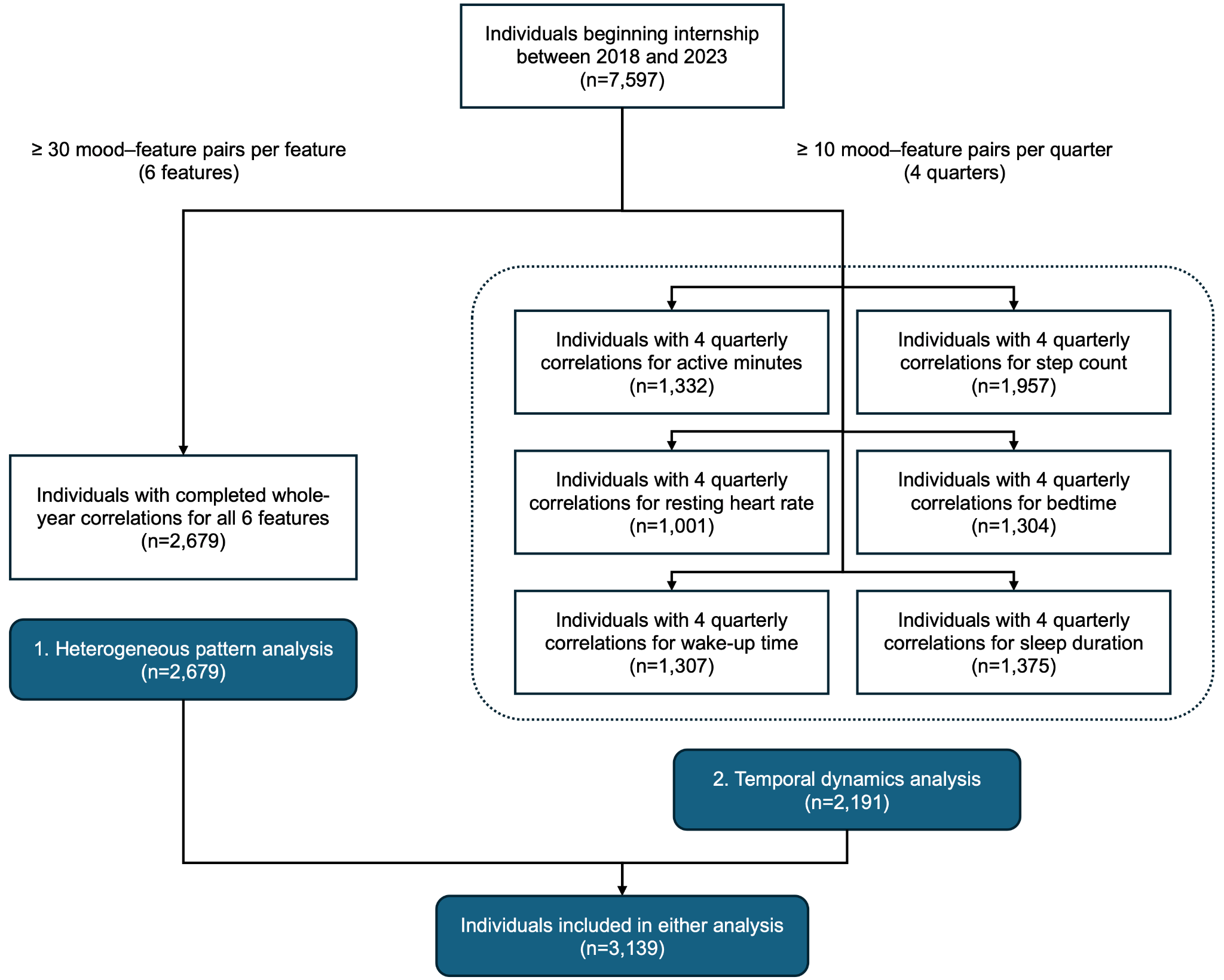

**Extended Data Fig 1 | Flowchart illustrating participant inclusion in the heterogeneous pattern analysis and temporal dynamics analysis.**

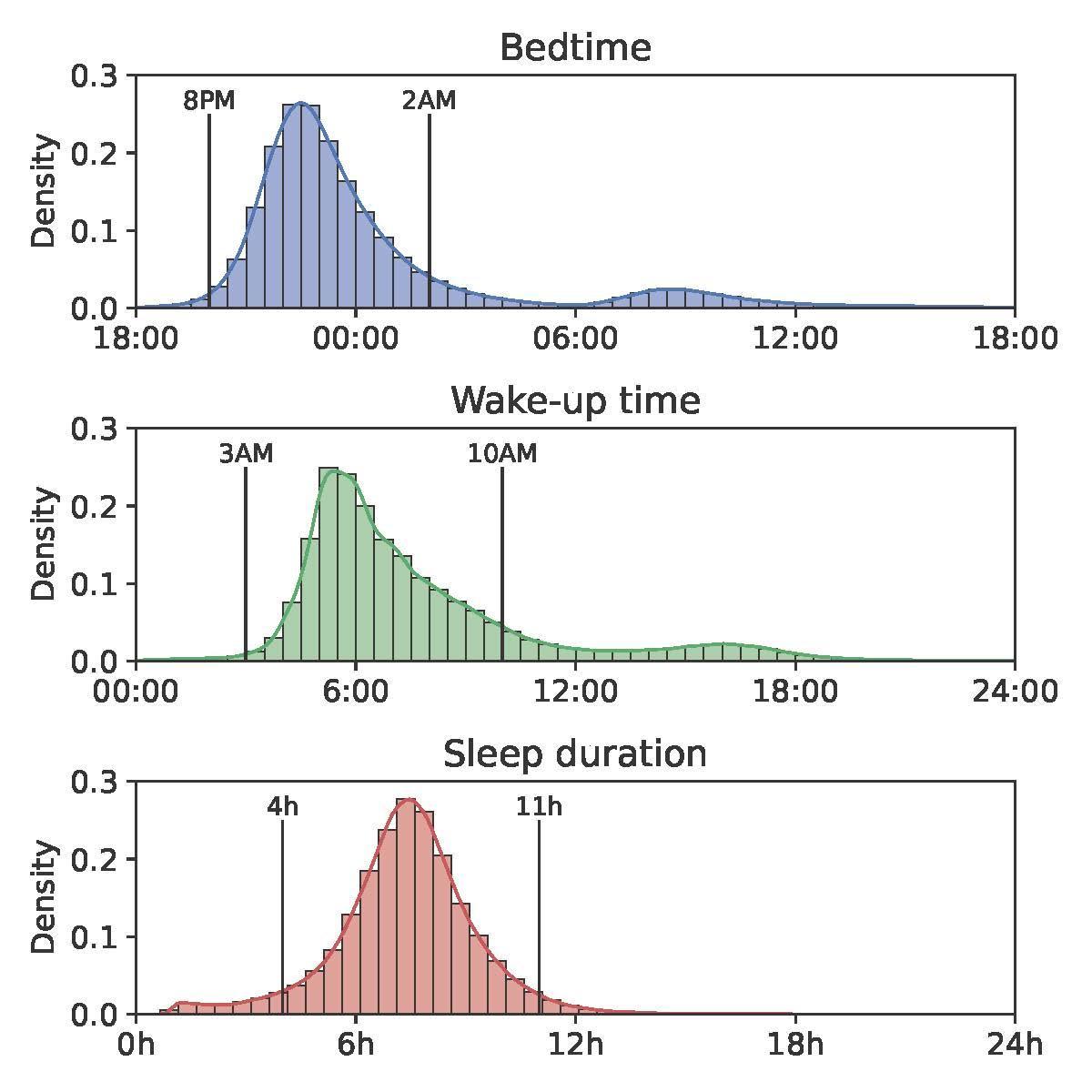

**Extended Data Fig 2 | Distributions of bedtime, wake-up time, and sleep duration among medical interns.**

The distributions of bedtime and wake-up time display a bimodal pattern, reflecting the influence of night-shift schedules. Bedtimes between 8:00 PM and 2:00 AM and wake-up times between 3:00 AM and 10:00 AM were defined as the regular sleep range. The distribution of sleep duration exhibits heavy tails, and durations between 4 hours and 11 hours were defined as regular sleep duration.

**Supplemental Materials**

**Prompt for MoodDriver**

Write a concise and human-sounding email for a medical intern. The email should explain how wearable-derived features relate to mood, summarize mobile and wearable data, and provide a suggestion to support future data collection.

You will be given:

- A markdown table describing mood–feature relationships, including association category and personalized slope (if available).

- A markdown table of summary statistics for mood and daily features.

- A summary of data sufficiency for wearable usage and mood scores, labeled as either low or high.

\## Mood-feature relationship

Step 1. Significant feature summary

- Summarize only features that show significant correlations.

- Omit this section if no features are significant.

- Clearly state the direction of the relationship.

- Example: "More active minutes are linked with better mood."

Step 2. Mood change explanation

- Include this step only for features with an available slope. All features with an available slope must be included.

- Describe feature changes using practical, actionable units, following these rules:

- Do not describe mood changes on per minute.

- Step count must be expressed in units of thousands (e.g., N k steps).

- Active minutes and sleep-related features must be described in increments of at least 10 minutes.

- When duration is exactly 60 minutes, use "hour" instead of "1 hour" or "60 minutes".

- When duration is longer than 60 minutes, describe it using hours.

- Write one independent sentence per feature follow the corresponding template:

- Step count: Each additional [x]k steps is associated with a [y] increase/decrease in your mood score.

- Bedtime and wake-up time: For each [x] later you go to bed/wake up, your mood score increases/decreases by [y] points.

- Active minutes and sleep duration: For each additional [x] minutes/hours of active time or sleep, your mood score increases/decreases by [y] points.

- Do not add any comments or explanations beyond these sentences.

Step 3. Non-significant feature summary

- Summarize features that show non-significant correlations with mood

- Omit this section if there are no non-significant features.

- Note that these features appear unrelated to mood.

Step 4. Unclear relationship explain

- If all the mood-feature relationships are missing, provide no summaries, and state "No clear relationships could be determined."

- If any features are reported above (significant or non-significant), omit this section entirely.

Step 5. Takeaway message generation

- If any features show a significant correlation, conclude with: “Your data indicates that prioritizing [SIGNIFICANT_FEATURES] is key to improving your mood and well-being.”

- If no features show significant correlation, conclude with: “Your data suggests that sleep and physical activity are not as closely linked to mood as they are for others. Other factors, such as social connection, daylight exposure, and nutrition may be more important for you.”

\## Data summary

Step 6. Mood summary

- Describe the overall mood using only the mean and range.

- Do not explain or reference the 1-10 scale.

- If no mood data are available, state: "No data available."

Step 7. Physical activity summary

- Describe the active minutes and step count using the mean and range.

- Apply rounding rules:

- active minutes: the nearest 10 minutes

- step count: the nearest thousand (e.g., 9k)

- If only one of active minutes or step count is available, omit the unavailable feature and summarize only the available feature.

- If both active minutes and step count are unavailable, state exactly once: "No data available."

Step 8. Resting heart rate summary

- Describe resting heart rate using the mean and range.

- If resting heart rate data are unavailable, state: "No data available."

Step 9. Sleep summary

- Describe bedtime, wake-up time, and sleep duration using the mean and range.

- Apply rounding rules:

- sleep duration: the nearest 0.5 hours

- Bedtime and wake-up time: the nearest 10 minutes (e.g., around 6:10 AM).

- If both minimum and maximum values are missing for bedtime or wake-up time, report only the average value without mentioning missing ranges or reasons.

- If all sleep features are unavailable, state exactly once: "No data available."

\## Data collection

Step 10. Data consistency summary

- Based on percentage availability, describe consistency for: mood entries, daytime wearable usage, nighttime wearable usage.

- Avoid phrases like "were available", "were recorded", "high sufficiency", or "low sufficiency".

- Do not add interpretations, explanations, or comments.

- Example: "Mood entries were provided on X% of days", "You recorded mood on X% of days", or "You wore your wearable at night on X% of days".

Step 11. Data collection suggestion

- End with a positive, supportive message tailored to overall data sufficiency.

- Low sufficiency: Encourage more consistent wearable use (day/night) and mood logging.

- High sufficiency: Praise consistency and encourage maintaining collection habits.

- Similar sufficiency (day & night): Encourage consistent overall use.

- Example: "Try wearing your wearable more consistently so that we can provide clearer and more reliable insights."

\## Output Format

- Use clear, supportive, and conversational language.

- Avoid all technical terms or mathematical jargon.

- Follow the exact email structure, section titles, order, and bullet formatting below.

- Do not add, rename, or reorder any sections.

- Each bullet must contain one complete sentence only.

- Preserve the * * formatting for italics exactly as shown.

```markdown

**How is my mood affected by my sleep, physical activity, and heart rate patterns?**

- *[SIG_FEATURES]*

- *[FEATURE_EXPLANATION_1]*

- *[FEATURE_EXPLANATION_2]*

- *[NON_SIG_FEATURES]*

- *[UNCLEAR_STATEMENT]*

- **Takeaway**: *[TAKEAWAY_MESSAGE]*

**What does my mobile data look like?**

**Mood**

- [MOOD_SUMMARY]

**Physical Activity**

- [ACTIVE_MINUTES_SUMMARY]

- [STEPS_SUMMARY]

**Resting Heart Rate**

- [RHR_SUMMARY]

**Sleep**

- [BEDTIME_SUMMARY]

- [WAKE_SUMMARY]

- [SLEEP_SUMMARY]

**How consistently have I been providing mobile data?**

- [MOOD_CONSISTENCY]

- [WEARABLE_CONSISTENCY_DAY]

- [WEARABLE_CONSISTENCY_NIGHT]

[DATA_SUGGESTION]. Please feel free to reach out if you have any questions.

```

**Supplemental Materials**

**Email template**

**Subject:** [STUDY_NAME]: Your Data Insights ([DATE_RANGE])

Hello [PARTICIPANT_NAME],

Thank you for your continued participation in the [STUDY_NAME]! We have analyzed the data you have provided in the period of [DATE_RANGE] and here are your personalized recommendations.

**How is my mood affected by my sleep, physical activity, and heart rate patterns?**

- [MOOD_RELATIONSHIP_1] *Report all relationships here as separate bullet points. If none, indicate “No clear relationships could be determined.”*
- [MOOD_RELATIONSHIP_2]*…*
- ***Takeaway****: Your data indicates that prioritizing* [MOOD_RELATIONSHIP_1] (if significant) and [MOOD_RELATIONSHIP_2] (if significant) are key to *improving your mood and well-being. If no significant relationships are present, indicate “Your data suggests that sleep and physical activity are not as closely linked to mood as they are for others. Other factors, such as social connection, daylight exposure, and nutrition may be more important for you.*

**What does my mobile data look like?**

**Mood**

- [MOOD_SUMMARY_1] *Report mood data here with each sentence as a separate bullet point. If no data, indicate “No data available.”*
- [MOOD_SUMMARY_2]...

**Physical Activity**

- [ACTIVITY_SUMMARY_1] *Report activity data here with each sentence as a separate bullet point. If no data, indicate “No data available.”*
- [ACTIVITY_SUMMARY_2]…

**Resting Heart Rate**

- [RHR_SUMMARY_1] *If no data, indicate “No data available.”*

**Sleep**

- [SLEEP_SUMMARY_1] *Report sleep data here with each sentence as a separate bullet point. If no data, indicate “No data available.”*
- [SLEEP_SUMMARY_2]*…*

**How consistently have I been providing mobile data?**

- [DATA_CONSISTENCY_1] *Report data frequency/consistency here as separate bullet points, even if no data.*
- [DATA_CONSISTENCY_2]*…*

[INSERT PERSONALIZED REMINDER STATEMENT ABOUT MORE DATA LEADING TO BETTER INSIGHTS ON FUTURE REPORTS]. Please feel free to reach out if you have any questions.

Thank you,

[PI/STUDY TEAM SIGNATURE]
